# Interaction between malarial transmission and BCG vaccination with COVID-19 incidence in the world map: A cross-sectional study

**DOI:** 10.1101/2020.04.03.20052563

**Authors:** Rudra Prosad Goswami, Dheeraj Mittal, Rama Prosad Goswami

**Author notes:** **Corresponding Author:** Rudra Prosad Goswami, DM, Assistant Professor, Department of Rheumatology, All India Institute of Medical Sciences, New Delhi, India.

## Abstract

**Background:** COVID-19 (Corona virus Disease-2019) is a new public health emergency and is a pandemic currently. Incidence and mortality of COVID-19 vary in different geographical areas. In this study we aimed to analyse the relationship between malaria transmission and BCG vaccination with COVID-19 incidence in the world map.

**Materials and methods:** We collected malaria cases data (World Health Organisation (WHO), 2018), worldwide COVID-19 cases and mortality data (European Centre for Disease Prevention and Control) and data on BCG vaccination. COVID-19 incidence and mortality was compared.

**Findings:** Data on 5316978938 persons from 166 countries were analysed. Malaria incidence rate was negatively correlated with COVID-19 incidence rate (correlation coefficient = -0.513, p<0.001). Malaria free countries had significantly higher number of COVID-19 cases compared to malaria endemic countries. In Europe and Americas, countries, which have higher BCG vaccination coverage, had significantly less mortality per thousand population compared to those with low BCG coverage (median 0.0002 (0-0.0005) vs 0.0029 (0.0002-0.0177), p=0.017). The case fatality ratio of COVID-19 was related nonlinearly to the malaria incidence.

**Conclusions:** The results suggest the changing human immune system as we progress to eliminate parasitic diseases with time. Chloroquine exposure in malaria endemic zones might have a protective effect.

**Summary box:** *“What is already known on this subject?”:* To the best of the authors no similar evidence, of the effect of malarial transmission on the COVID-19 global distribution is known. The effect of the Bacille Calmette Guérin (BCG) vaccine on modifying the human immune system has been reported before and is postulated to protective against certain viral infections like Influenza A (H1N1) and herpes virus.

*“What this study adds?”:* This study finds that COVID-19 incidence, worldwide is less in countries, which are malaria-endemic. In the European and American countries, increased BCG coverage may have some mortality advantage against COVID-19. The case fatality rate was related to malaria incidence, however, in a complex way. This could be a window into the changing landscape of human immune system as we progress to eliminate parasitic disease with time or this could be due to long-term protective body level of anti-malarials like chloroquine or hydroxychloroquine in countries with higher malaria incidence rate.

## Introduction

The COVID-19 (Coronavirus Disease-2019) is a public health emergency of international concern and currently is a pandemic. Approximately 15% of patients suffer from severe form of the disease.^1^ As of this time there is no known specific, effective and evidence based treatment.

We noted that COVID-19 transmission and mortality are apparently higher in the United States and European countries in contrast to malaria and other infectious diseases endemic regions like South East Asian and African countries.

The Bacille Calmette Guérin (BCG) vaccine is a live attenuated vaccine against tuberculosis and apart from providing some protection against development of severe forms of tuberculosis it has been shown to have significant immunomodulatory action and protection against certain viral infections like respiratory syncytial virus, Influenza A (H1N1) and herpes virus.^2^

The interaction between exposure to malaria infection and BCG vaccination with incidence of COVID-19 has not been explored.

So, in the absence of any such data, we aimed to analyse the hypothesis regarding the relationship between malaria incidence and BCG coverage with COVID-19 incidence in the world map.

## Materials and methods

For this cross sectional study, we collected malaria cases and mortality data (2018) from the World Health Organisation (WHO),^3^ and worldwide COVID-19 cases and mortality data from the European Centre for Disease Prevention and Control website.^4^ Data on BCG vaccination coverage was accessed from the WHO website and other online sources.^5, 6^ Population data was confirmed from the world population website.^7^ Only autochthonous malaria cases were considered and migrant cases were not considered. All data were compiled on an excel sheet and then transferred to SPSS ver. 21.

### Statistical analysis

Malaria and COVID-19 incidence rate was calculated as per thousand of population. Deviation from normal distribution was tested with Shapiro-Wilk test. All continuous data were expressed as mean ± standard deviation (SD) or median (interquartile range, IQR) as appropriate and discrete data as fraction (percentage). As a first step non-parametric correlation (Kendall’s Tau) was calculated between malaria incidence and mortality with COVID-19 incidence. Then COVID-19 incidence was compared between countries, which are malaria free or not. Similar comparisons were made with BCG vaccination coverage. Mortality of COVID-19 was assessed as per thousand-population in each country. Association of mortality from COVID-19 was tested against malaria incidence and BCG coverage. To correct for false inflation of mortality rate, mortality analysis was restricted to only those countries reporting more than 100 COVID-19 cases. Comparison of means was done with Mann Whitney U test. A binary logistic regression was run with COVID-19 incidence as dependent variable and the following two variables: malaria free country and BCG vaccination coverage ≤95%. Analysis of incidence and mortality of COVID-19 with malaria incidence and BCG coverage was done separately for different geographical location of the countries. Finally, case fatality rate of COVID-19 was analysed in comparison to malaria incidence with non-linear regression.

## Results

Overall data on 5316978938 persons from 166 countries were extracted. Number of malaria cases was 228435738 and number of malaria deaths was 405255. Number of malaria free countries was 81. Number of COVID-19 cases was 412637 and number of COVID-19 deaths was 18559. The median incidence of COVID-19 per thousand populations per country was 0.009 (IQR: 0.0007-0.12) and the median mortality per thousand populations per country was 0.0003 (IQR: 0-0.0018). The COVID-19 incidence was highest in the European/American countries (0.0974, IQR: 0.024-0.33) followed by Asian/Australasian countries (0.0084, IQR: 0.0005-0.077) followed by African countries (0.0015, IQR: 0.0002-0.0045) and the difference was statistically significant (p<0.001). Median case fatality rate of COVID-19 was 1.68% (IQR: 0.39-6.5).

Data on BCG vaccination coverage was available from 148 countries. Number of countries achieving BCG vaccination coverage of >95% was 53 (31.9%). BCG vaccination coverage of >95% was present in 40.3% of European or American countries (29/72), 25% of African countries (11/44) and 40.6% of Asian/Australasian countries (13/32).

Median malaria cases per 1000 population were 0.01 (IQR: 0 to 33). Number of malaria free countries was 81 (48.8%). Number of malaria free countries was significantly higher in the European/American continents (75.9%, 66/87) compared to the African (2.3%, 1/44) or Asian/Australasian (40%, 14/35) countries (p<0.001).

Malaria incidence rate was negatively correlated with COVID-19 incidence rate (tau = -0.513, p<0.001) Malaria mortality rare was also negatively correlated with COVID-19 incidence rate (tau= -0.335, p<0.001).

Both total number of COVID-19 cases (7 (IQR 2-89) vs. 249 (86-1492) respectively, p<0.001) and COVID-19 incidence (0.0008 (IQR 0.0001-0.006) vs. 0.09 (IQR: 0.02-0.34) respectively, p<0.001) was lower in countries where malaria occurred versus malaria free countries. Although COVID-19 incidence (0.02 (IQR 0.003-0.11) vs. (IQR: 0.0002-0.06) respectively, p=0.01) was significantly higher in countries with >95% BCG coverage compared to countries with ≤95% BCG coverage, total number of COVID-19 cases per country were similar (102 (IQR: 26-469) vs 42 (IQR 3-562) p=0.25). The COVID-19 mortality rate was higher in malaria free countries (Table 1).

**Table 1.** Comparison of nCOVID-19 case and incidence rate between different groups of countries based on malaria and BCG coverage

| **Parameters** | **Malaria free**<br>**n=81** | **Not malaria free**<br>**n=85** | **p-value** | **BCG vaccination**<br>**coverage ≤95%**<br>**n=95** | **BCG vaccination**<br>**coverage >95%**<br>**n=53** | **P - value** |
| --- | --- | --- | --- | --- | --- | --- |
| nCOVID-19 cases | 4830 (14402) | 250 (1030) | <0.001 | 2495 (9782) | 2500 (11673) | 0.25 |
| nCOVID-19 incidence (per thousand) | 0.75 (3.3) | 0.07 (0.36) | <0.001 | 0.13 (0.37) | 0.72 (4.08) | 0.01 |
| **Parameters** | **Malaria free**<br>**n=80** | **Not malaria free**<br>**n=44** | **p-value** | **BCG vaccination**<br>**coverage ≤95%**<br>**n=64** | **BCG vaccination**<br>**coverage >95%**<br>**n=44** | **P - value** |
| nCOVID-19 mortality (per thousand) | 0.04 (0.26) | 0.002 (0.009) | 0.03 | 0.0049 (0.02) | 0.05 (0.35) | 0.93 |

Cells are expressed as mean (standard deviation)
All statistical significance testing were done with the Mann Whitney U test

In multivariate binary logistic regression (Table 2) the chance of encountering higher number of COVID-19 cases was higher in malaria free countries (OR: 16.76, 95% CI: 7.3-38.5, p<0.001) only.

**Table 2.** Results from the binary logistic regression with nCOVID-19 incidence as the dependent variable.

| **Factors** | **B** | **SE** | **OR** | **95% CI** | **P - value** |
| --- | --- | --- | --- | --- | --- |
| BCG vaccination coverage $\leq 95\%$ | -0.74 | 0.44 | 0.47 | 0.2-1.12 | 0.09 |
| Malaria free country | 2.81 | 0.42 | 16.76 | 7.3-38.5 | <0.000001 |

Abbreviations:
B: beta coefficient; SE: standard error; OR: Odd's ratio; CI: confidence interval
Hosmer Lemeshow Test: Chi-squared: 4.46, df: 2; p=0.11
Nagelkarke R squared: 0.47
Collinearity diagnostics: Variance inflation factor: malaria free country: 1.046; BCG vaccination coverage: 1.046

Next we analysed the effect of BCG coverage and malaria incidence with respect to geographic location of the countries (Table 3). In European and American countries, countries with >95% BCG coverage had lower COVID-19 mortality compared to countries with ≤95% coverage (respectively, 0.0002 (IQR 0-0.0005) vs 0.0029 (IQR: 0.0002-0.0177), p=0.017). However in African, Asian and Australasian countries increased BCG coverage was associated with increased COVID-19 incidence and numerically increased mortality. In all geographic locales malaria-free countries had higher COVID-19 incidence.

**Table 3.** Comparison of nCOVID-19 incidence and mortality according to BCG coverage according geographic locations.

| **European and American countries** |  |  |  |  |  |  |
| --- | --- | --- | --- | --- | --- | --- |
| Parameters | Malaria free<br>n=66 | Not malaria free<br>n=21 | p-value | BCG coverage ≤95%<br>n=43 | BCG coverage >95%<br>n=29 | P - value |
| nCOVID-19 incidence (per thousand) | 0.45 (0.86) | 0.13 (0.46) | 0.0001 | 0.29 (0.51) | 0.14 (0.33) | 0.28 |
| | Malaria free<br>n=48 | Not malaria free<br>n=8 | p-value | BCG coverage ≤95%<br>n=30 | BCG coverage >95%<br>n=17 | P - value |
| nCOVID-19 mortality (per thousand) | 0.019 (0.09) | 0.007 (0.02) | 0.7 | 0.009 (0.02) | 0.001 (0.002) | 0.017 |
| **African and Asia and Australasian countries** |  |  |  |  |  |  |
| Parameters | Malaria free<br>n=15 | Not malaria free<br>n=64 | p-value | BCG coverage ≤95%<br>n=52 | BCG coverage >95%<br>n=24 | P - value |
| nCOVID-19 incidence (per thousand) | 2.11 (7.63) | 0.05 (0.31) | 0.00001 | 0.006 (0.02) | 1.42 (6.05) | 0.0008 |
| | Malaria free<br>n=12 | Not malaria free<br>n=9 | p-value | BCG coverage ≤95%<br>n=8 | BCG coverage >95%<br>n=10 | P - value |
| nCOVID-19 mortality (per thousand) | 0.19 (0.66) | 0.004 (0.01) | 0.15 | 0.0008 (0.0001) | 0.23 (0.73) | 0.068 |

Cell entries indicate mean (standard deviation)
All statistical significance testing were done with the Mann Whitney U test

In non-linear regression of COVID-19 case fatality was compared to malaria incidence in each countries and a quartic equation (R squared=0.527) was the best fit as follows: COVID-19 case fatality = -0.033 + 7.9*(malaria incidence) -0.709*(malaria incidence)^2 + 0.022*(malaria incidence)^3-0.0002*(malaria incidence)^4. Linear regression with transformed malaria incidence as independent variable showed significant association with COVID-19 case fatality (B=0.235, 95% CI: 0.026-0.444, p=0.029, variance inflation factor=1.0).

## Discussion

The COVID-19 is a new pandemic. We empirically observed that malaria free countries have a higher incidence of COVID-19 among different countries. Currently we can offer no exact explanation of our findings. Such strong statistical association as observed is unlikely to be a chance association. Malarial transmission may give some protection against COVID-19. One explanation may be community level use of drugs such as chloroquine or hydroxychloroquine in malaria-endemic countries and relative non-use of them in malaria-free countries. Interestingly chloroquine has been shown to be effective in reduction of viral replications including COVID-19.^8,9^ Hydroxychloroquine has some immunomodulatory action and after the outbreak, studies have shown some activity of this drug.^10^ Viral inhibition from possibly higher blood level in malaria-endemic countries is one postulation. The second possible mechanism seems to be immunological. With increasing effort to eliminate and finally eradicate malaria long-term changes may occur in the human immune system. Possible link between filarial elimination and increased autoimmunity has previously been reported.^11^ Whether elimination of parasites leads to permanent changes in the human immune system should be investigated.

Prior BCG vaccination had an apparent paradoxical effect. In the overall analysis lower BCG coverage was associated with lower number of COVID-19 cases. However this effect is probably related to geographic locale of the countries. In European and American countries, where natural tubercular infection rate was low, higher BCG coverage was associated with decreased COVID-19 mortality. In these countries higher BCG coverage was also associated with numerically lower COVID-19 incidence. On the other hand, in the African, Asian and Australasian continent, where natural TB incidence is higher, increased BCG coverage appeared to have facilitated COVID-19 infection spread and mortality. Whether, after adequate control of spread of tuberculosis, the immunmodulatory effect of continued BCG vaccination, may help the human immune system to thwart future viral epidemics like the current COVID-19, is an open and interesting question.

The relationship of case fatality rate of COVID-19 with malaria incidence was complex and a quartic equation was needed for adequate modeling. In the equation the even powers have negative weights and the odd powers have positive weights. Thus in areas of non-zero low to very low malaria incidence, the effect of higher powers shall be overwhelmed by the linear terms and an increased COVID-19 case fatality is seen. On the other hand in malaria-endemic countries the reverse is observed.

There are some limitations in this study. Many other parasitic diseases could have been screened, which however due to lack of worldwide data could not be done. Many factors like underreporting of cases might underwhelm this report. Socioeconomics is an important factor, which is missing from the current analysis. To summarize, malaria free countries, worldwide, are experiencing higher burden of COVID-19 pandemic. Among the western countries, those, which have continued high BCG vaccination coverage, have an apparent mortality benefit from COVID-19 infection. However, among the African, Asian and Australasian countries, continued high-level BCG vaccination was associated with increased incidence of and mortality from the COVID-19 infection. From the COVID-19 case fatality analysis a lower mortality could be expected in malaria-endemic countries.

## Data Availability

With the first author....available on online repositories and available on request

